# Prognostic Value of Myocardial Perfusion Imaging by Cadmium Zinc Telluride Single-photon Emission Computed Tomography in Patients with Suspected or Known Coronary Artery Disease A Systematic Review and Meta-analysis

**DOI:** 10.1101/2023.05.10.23289824

**Authors:** Roberta Assante, Emilia Zampella, Valeria Cantoni, Roberta Green, Adriana D’Antonio, Teresa Mannarino, Valeria Gaudieri, Carmela Nappi, Pietro Buongiorno, Mariarosaria Panico, Mario Petretta, Alberto Cuocolo, Wanda Acampa

## Abstract

**Background:** Aim of this study was to define the prognostic value of stress myocardial perfusion imaging by cadmium zinc telluride (CZT) single-photon emission computed tomography (SPECT) for prediction of adverse cardiovascular events in patients with known or suspected coronary artery disease (CAD).

**Methods and Results:** Studies published until November 2022 were identified by database search. We included studies using stress myocardial perfusion imaging by CZT-SPECT to evaluate subjects with known or suspected CAD and providing primary data of adverse cardiovascular events. Total of 12 studies were finally included recruiting 36,415 patients. Pooled hazard ratio (HR) for the occurrence of adverse events was 2.17 (95% confidence interval, CI, 1.78-2.65) and heterogeneity was 66.1% (*P*=0.001). Five studies reported data on adjusted HR for the occurrence of adverse events. Pooled HR was 1.69 (95% CI, 1.44-1.98) and heterogeneity was 44.9% (*P*=0.123). Seven studies reported data on unadjusted HR for the occurrence of adverse events. Pooled HR was 2.72 (95% CI, 2.00-3.70). Nine studies reported data useful to calculate separately the incidence rate of adverse events in patients with abnormal and normal myocardial perfusion. Pooled incidence rate ratio was 2.38 (95% CI, 1.39-4.06) and heterogeneity was 84.6% (*P*<0.001). The funnel plot showed no evidence of asymmetry (*P*=0.517). At meta-regression analysis, we found an association between HR for adverse events and presence of angina symptoms and family history of CAD.

**Conclusions:** Stress myocardial perfusion imaging by CZT-SPECT is a valuable noninvasive prognostic indicator for adverse cardiovascular events in patients with known or suspected CAD.

## CLINICAL PERSPECTIVE

Stress single-photon emission computed tomography represents the most widely used non-invasive cardiac imaging tool for risk stratification of patients with suspected or known coronary artery disease. In the last decades, dedicated gamma cameras with semiconductor cadmium-zinc-telluride detectors for cardiac imaging resulted in improvement of both image accuracy and acquisition protocol of radionuclide perfusion imaging, followed by an optimal diagnostic and prognostic accuracy. Despite cadmium-zinc-telluride scanners have been introduced in the first decade of 21st century and the wide diffusion in the clinical practice, data full validating its value in risk stratification are not fully addressed. Based on data from 36,415 patients derived from 12 independent cohorts, we showed that the presence of an abnormal findings conferred about 2-fold increased risk of cardiovascular events. However, a high heterogeneity was observed among studies, ascribable to differences in stress test used, acquisition protocol, outcome measure and method used. Moreover, the majority of the selected studies in this metanalysis tested abnormality by summed stress score in a short-term follow-up. From a clinical perspective further studies should be provided, also in a long-term follow-up perspective, outlining the important role of other variables, as the extent and severity of myocardial ischemia, by cadmium-zinc-telluride for clinical decision making and prognostic evaluation addressing usage promotion decisions.

Stress single-photon emission computed tomography (SPECT) myocardial perfusion imaging represents the most widely used non-invasive cardiac imaging tool for diagnosis and risk stratification of patients with suspected or known coronary artery disease (CAD).^1^ The wide diffusion in the clinical practice of dedicated gamma cameras with semiconductor cadmium-zinc-telluride (CZT) detectors for cardiac imaging have provided an increase in photon sensitivity and in spatial resolution compared to the conventional SPECT system, leading to an improvement of both image accuracy and acquisition protocol.^2–7^ A recent meta-analysis showed that dynamic CZT-SPECT myocardial perfusion imaging has a good sensitivity and specificity in diagnosing CAD as compared to the gold standards used in clinical practice.^8^ Moreover, published data have been provided on the prognostic value of stress-rest myocardial perfusion imaging using CZT-SPECT showing similar prognostic results compared to conventional SPECT, with lower prevalence of hard events in patients with normal scan.^9–11^ However, despite CZT-SPECT scanner has been introduced in the first decade of 21st century,^12^ data addressing the prognostic value of CZT-SPECT using the different parameters of myocardial perfusion and function by CZT technology seem to be limited. To address this, we performed a systematic review and meta-analysis of the available observational studies to evaluate the prognostic value of CZT-SPECT in patients with suspected or known CAD.

## METHODS

This systematic review and meta-analysis followed the Preferred Reporting Items for Systematic Reviews and Meta-analyses (PRISMA) statement (see the Supplementary Material for PRISMA Checklist)^13^ and registered as record ID 417979 in the PROSPERO database (University of York, UK; http://www.crd.york.ac.uk/PROSPERO/).

### Data Sources and Study Selection

An English literature search was performed using the PubMed and Embase databases to identify articles published from inception until November 2022. Studies search was restricted to data obtained in humans and adults and was conducted using the following key words: “prognosis, prognostic value, clinical outcome, risk stratification, adverse outcome, follow-up, hazard ratio (OR HR), myocardial perfusion imaging (OR MPI), myocardial perfusion scintigraphy (OR MPS), single photon emission computed tomography (or SPECT), cadmium zinc telluride camera (OR CZT), coronary artery disease (OR CAD)”. The full search strategy is shown in the supplementary material. The title and abstract of potentially relevant studies were screened for appropriateness before retrieval of the full article by two reviewers (VC and RG), and disagreements were resolved by consensus. The full-published reports of the abstracts selected by the reviewers were retrieved and the same reviewers independently performed a second-step selection based on the eligibility criteria; disagreements were resolved by consensus. In addition, the bibliographies of retrieved articles were manually reviewed for additional citations. We included as supplementary material a list of full-text articles excluded and a table reporting the exclusion criteria for each article (Table S1).

### Study Eligibility and Data Extraction

Each study was initially identified considering journal, author, and year of publication. To harmonize the predictors of interest, a study was considered eligible if all of the following criteria were met: 1) the study presented data of subjects with known or suspected CAD referred for stress myocardial perfusion imaging by a CZT camera; 2) the study included at least 100 subjects; 3) the study provided unadjusted and/or adjusted HR of abnormal versus normal myocardial perfusion as dichotomous variable for the occurrence of adverse cardiovascular events; or the unadjusted and/or adjusted HR could be obtained from Kaplan-Meier curves; or the unadjusted and/or adjusted HR could be convert from three categories to two categories of abnormal versus normal perfusion. In case of multiple studies reported from the same research group, potential cohort duplication was avoided by including the largest study only. Population data were also collected on age and on prevalence of female sex, traditional cardiovascular risk factors (diabetes, dyslipidemia, smoking, hypertension, family history of CAD), angina-like symptoms, and history of CAD (including previous myocardial infarction and coronary revascularization). Follow-up time and occurrence of the different endpoints were recorded.

### Quality Assessment

We utilized the modified Quality in Prognostic Studies (QUIPS) appraisal tool considering outcome measurement, prognostic factor measurement, statistical analysis, study attrition, study confounding and study participation.^14^ Each domain contains several signal questions used to help judge the risk of bias (low, high, or unclear). First, the risk of bias was determined for each domain. Then, the overall risk for each study was judged. Study quality was not considered restrictive for inclusion, but it was comprehensively evaluated. Two reviewers (VC and RG) completed the screening process independently. Disagreement in the process of answering questions was discussed until consensus was reached.^14^

### Statistical Analysis

Continuous variables are expressed as mean ± SD and categorical data as percentages. To quantify the prognostic impact of abnormal vs. normal myocardial perfusion imaging, we chose as effect size the summary hazard ratio (HR) of abnormal myocardial perfusion findings across studies as previously described.^15^ The HR has a straightforward interpretation when translated to probability according to the formula: probability = (HR)/(1+HR).^16^ Indeed, a hazard ratio of 2 corresponds to a 67% chance for a randomly selected patient with abnormal perfusion to experience an event before a randomly selected patients with normal perfusion, and a hazard ratio of 3 corresponds to a 75% chance. We also calculated the incidence rate ratio (IRR) for the occurrence of adverse events in patients with abnormal and normal perfusion. The HR with 95% confidence interval (CI) of each study was abstracted.^17^ To minimize the effect of confounding, we included the most extensively adjusted HR (with associated 95% CI derived from multivariable regression analysis) from each original study, if available. For studies that did not provide adjusted HR, the univariable risk estimate was included in the analysis.

The main outcome of interest was the occurrence of adverse events, including death, myocardial infarction, late revascularization, stroke, heart failure or acute coronary syndrome. When available, a secondary analysis was performed considering only the occurrence of hard cardiac events (i.e., cardiac death and myocardial infarction). The summary estimates of HR and 95% CI were computed using the random effects model of DerSimonian and Laird.^18^ The HR were log transformed, to obtaining a sampling distribution closer to normality, and pooled using a random effects model. The estimates were back transformed, and results presented as HR. The weight of each study was calculated with the inverse variance method.^19^ Between-study heterogeneity was evaluated with Cochran’s Q and I^2^ statistics.^20^ When statistical heterogeneity was substantial, meta-regression analysis was performed to assess if study-level variables such as age, women, diabetes, dyslipidemia, smoking, hypertension, angina, family history of CAD, prior myocardial infarction, prior revascularization procedures and known CAD, were associated with HR. Publication bias, i.e. the likelihood that a published study will be influenced by its results, was graphically examined by the funnel plot obtained plotting for each study the ln HR versus its standard error, and also formally assessed with the regression test of asymmetry described by Egger et al.^21^ To adjust for funnel plot asymmetry, a trim and fill analysis was also performed.^22^ This procedure estimates the number and outcomes of theoretical missing studies and incorporates them into the meta-analysis.^23^ The basis of the method is to 1) trim (remove) the smaller studies causing funnel plot asymmetry, 2) use the trimmed funnel plot to estimate the true center of the funnel, then 3) replace the omitted studies and their missing counterparts around the center (filling). For summary HR, two subgroup analyses were also performed, one considering separately studies reporting adjusted or unadjusted HR, the other considering studies according to the camera technology used.

When it was feasible, for each study annualized event rates were calculated by dividing the number of events by the mean or median follow-up duration and the incidence rate ratio (IRR) with 95% CI were estimated to compare the outcome of patients with abnormal versus normal myocardial perfusion. As for HR, also the IRR were log transformed pooled using a random effects model and results presented as IRR after back-transformation. To evaluate the robustness of the results, a leave-one-out sensitivity analysis for IRR and HR for adverse events was performed by iteratively removing one study at a time to confirm that our findings were not driven by any single study.^24^ All analyses were performed using Stata, version 15.1 (StataCorp, College Station, TX). Two-sided *P* values ≤0.05 were considered statistically significant.

## RESULTS

### Search Results

The complete literature search is presented in Figure 1. The initial search identified 1,340 potentially eligible citations. The reviewers, after the evaluation of the titles and abstracts of these studies discharged 1,268 citations because they were judged to be non-relevant or non-pertinent. To determine eligibility, each investigator reviewed blinded the full text of the remaining 73 articles, and disagreements were resolved by consensus. After revision, 61 articles were excluded, remaining 12 articles including 36,415 patients.

**Figure 1.**
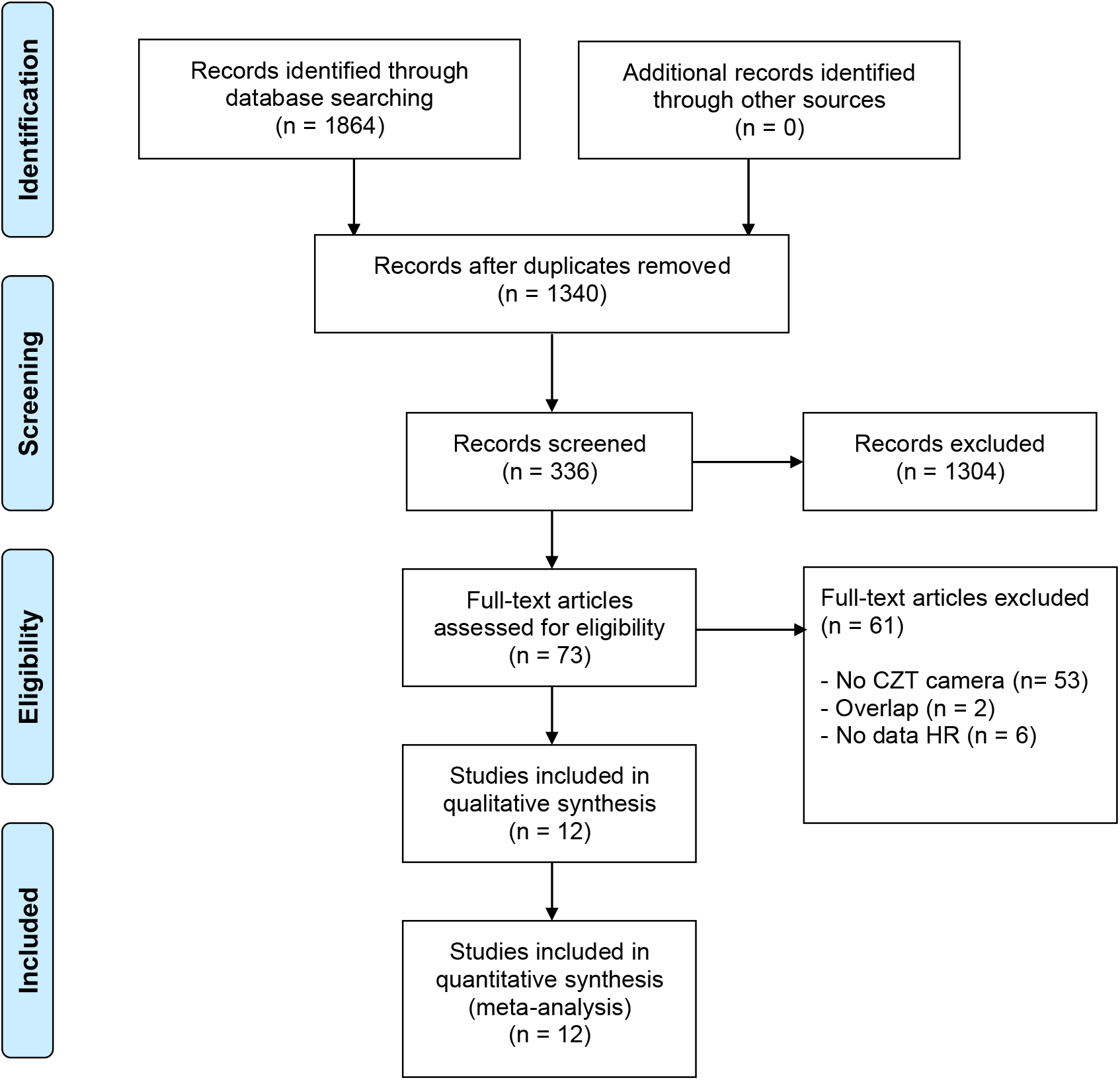
Literature search and selection process of studies included in analysis. HR, hazard ratio.

### Quality Assessment

Figure S1 summarizes the quality assessment of included studies using the QUIPS tool. The risk of bias was considered unclear overall. The domains that showed a sustained uncertain risk of bias were the ‘prognostic factor measurements’, ‘outcome measurement’ and “study confounding” due to the differences in stress test used, acquisition protocol, outcome measure and method used.

### Characteristics of Studies

Demographic data and clinical characteristics of patients are detailed in Table 1.^25–36^ Study sample size ranged from 151 to 19,495 subjects. For two studies,^31, 34^ we reported the number of patients with available CZT data. The mean age ranged from 62 to 69 years, with the proportion of women ranging from 20% to 59%. Follow-up ranged from 1.7 to 6.6 years. The characteristics of included studies are shown in Table 2.

**Table 1.**
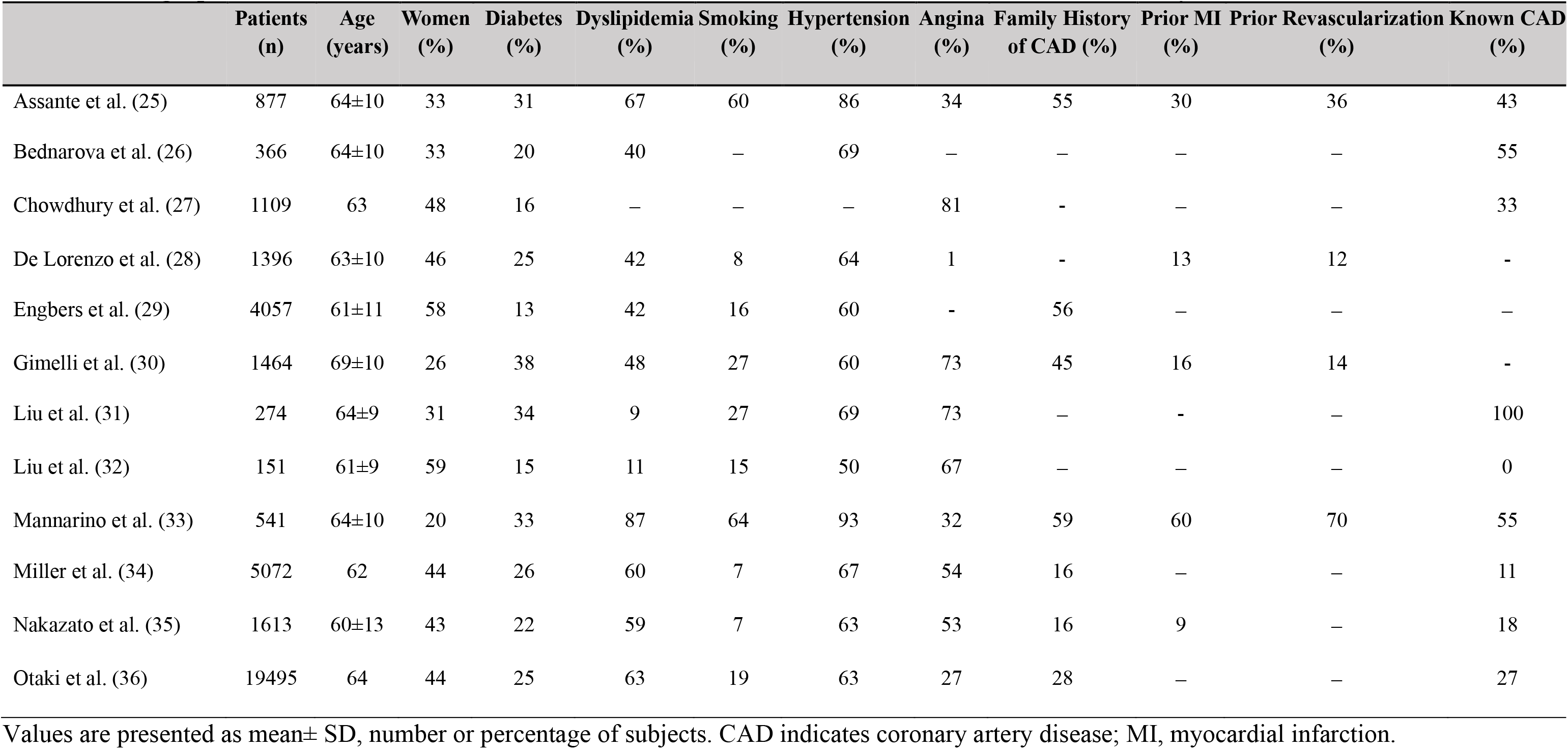
Demographic and Clinical Characteristics of Patients Included in the Studies Enrolled in Meta-analysis.

|  | Patients<br>(n) | Age<br>(years) | Women<br>(%) | Diabetes<br>(%) | Dyslipidemia<br>(%) | Smoking<br>(%) | Hypertension<br>(%) | Angina<br>(%) | Family History<br>of CAD (%) | Prior MI<br>(%) | Prior Revascularization<br>(%) | Known CAD<br>(%) |
| --- | --- | --- | --- | --- | --- | --- | --- | --- | --- | --- | --- | --- |
| Assante et al. (25) | 877 | 64±10 | 33 | 31 | 67 | 60 | 86 | 34 | 55 | 30 | 36 | 43 |
| Bednarova et al. (26) | 366 | 64±10 | 33 | 20 | 40 | – | 69 | – | – | – | – | 55 |
| Chowdhury et al. (27) | 1109 | 63 | 48 | 16 | – | – | – | 81 | - | – | – | 33 |
| De Lorenzo et al. (28) | 1396 | 63±10 | 46 | 25 | 42 | 8 | 64 | 1 | - | 13 | 12 | - |
| Engbers et al. (29) | 4057 | 61±11 | 58 | 13 | 42 | 16 | 60 | - | 56 | – | – | – |
| Gimelli et al. (30) | 1464 | 69±10 | 26 | 38 | 48 | 27 | 60 | 73 | 45 | 16 | 14 | - |
| Liu et al. (31) | 274 | 64±9 | 31 | 34 | 9 | 27 | 69 | 73 | – | - | – | 100 |
| Liu et al. (32) | 151 | 61±9 | 59 | 15 | 11 | 15 | 50 | 67 | – | – | – | 0 |
| Mannarino et al. (33) | 541 | 64±10 | 20 | 33 | 87 | 64 | 93 | 32 | 59 | 60 | 70 | 55 |
| Miller et al. (34) | 5072 | 62 | 44 | 26 | 60 | 7 | 67 | 54 | 16 | – | – | 11 |
| Nakazato et al. (35) | 1613 | 60±13 | 43 | 22 | 59 | 7 | 63 | 53 | 16 | 9 | – | 18 |
| Otaki et al. (36) | 19495 | 64 | 44 | 25 | 63 | 19 | 63 | 27 | 28 | – | – | 27 |

Values are presented as mean± SD, number or percentage of subjects. CAD indicates coronary artery disease; MI, myocardial infarction.

**Table 2.** Characteristics of Included Studies Reporting Adverse Events.

|  | Stress Test | End Point | Follow-up (years) | Abnormal Perfusion Criteria | Data in Meta-analysis | Multivariable Adjustment |
| --- | --- | --- | --- | --- | --- | --- |
| Assante et al. (25) | Exercise, dipyridamole | CD, MI, R | 2.2 | TPD $\geq 5$ % | Multivariate HR | Not adjusted |
| Bednarova et al. (26) | Exercise, regadenoson | CD, MI, UA, R, HF | 1.9 | Presence of ischemia | Multivariate HR | Not adjusted |
| Chowdhury et al. (27) | Adenosine | CD, MI | 1.7 | SSS $\geq 4$ | Univariate HR* | Not adjusted |
| De Lorenzo et al. (28) | Exercise, dipyridamole, dobutamine | ACD | 2.08 | SSS $> 4$ | Univariate HR | Clinical and imaging variables |
| Engbers et al. (29) | Exercise, pharmacologic | CD, MI | 2.4 | SDS $\geq 2$ | Multivariate HR** | Not adjusted |
| Gimelli et al. (30) | Exercise, dipyridamole | CD, MI | 3.5 | Best cut-off SSS $> 8$ | Multivariate HR | Not adjusted |
| Liu et al. (31) | Exercise, adenosine | CD, MI, UA, R, HF, Stroke | 2.9 | SSS $\geq 4$ in individual coronary territories | Multivariate HR | Number of events |
| Liu et al. (32) | Exercise, adenosine | CD, MI, UA, R, HF, Stroke | 2.9 | SSS $\geq 4$ in individual coronary territories | Multivariate HR | Number of events |
| Mannarino et al. (33) | Exercise, dipyridamole | CD, MI, R | 2.2 | SSS $> 3$ | Univariate HR* | Not adjusted |
| Miller et al. (34) | Exercise, pharmacologic | ACD | 6.6 | SSS $\geq 1$ | Multivariate HR** | Clinical and imaging variables |
| Nakazato et al. (35) | Exercise, adenosine | ACD | 2.6±0.5 | SSS $> 3$ | Multivariate HR** | Not adjusted |
| Otaki et al. (36) | Exercise, pharmacologic | ACD, MI, UA, R | 4.5±0.5 | Visual perfusion assessment | Multivariate HR | Clinical variables |

CD indicates cardiac death; MI, myocardial infarction; R, revascularization; UA, unstable angina; HF, heart failure; ACD, all-case death; TPD, total perfusion defect; SSS, summed stress score; SDS, summed difference score; HR, hazard ratio.
\*Derived from Kaplan-Meier analysis.
\*\*Converted from three categories.

### Prognostic Value of Stress Imaging

The HR for the occurrence of adverse events was reported in 12 studies. The HR ranged from 1.46 to 7.92. The pooled HR was 2.17 (95% CI, 1.78-2.65) and the heterogeneity was 66.1% (*P*=0.001) (Figure 2). Among the included publications, three studies reported the HR for the occurrence of hard events, and the HR ranged from 3.25 to 7.92. From these studies, the pooled HR was 3.39 (95% CI, 2.47-4.64) (Figure 3).

**Figure 2.**
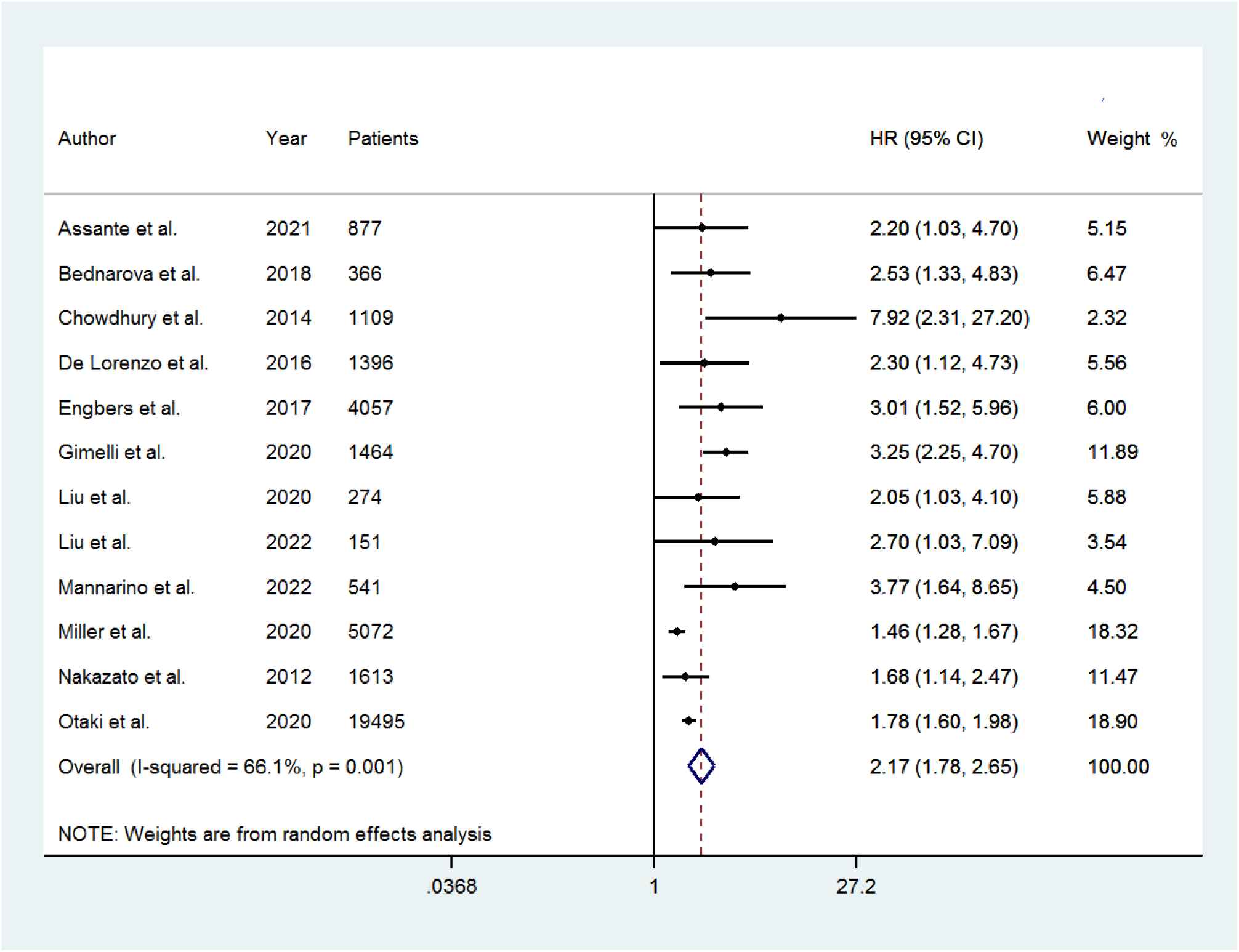
Forest plot of HR for adverse events associated with abnormal myocardial perfusion. Horizontal lines represent 95% CI of the point estimates. The diamond represents the pooled estimate (size of the diamond = 95% CI). The solid vertical line represents the reference of no increased risk, and the dashed vertical line represents the overall point estimate; HR, hazard ratio; CI, confidence interval.

**Figure 3.**
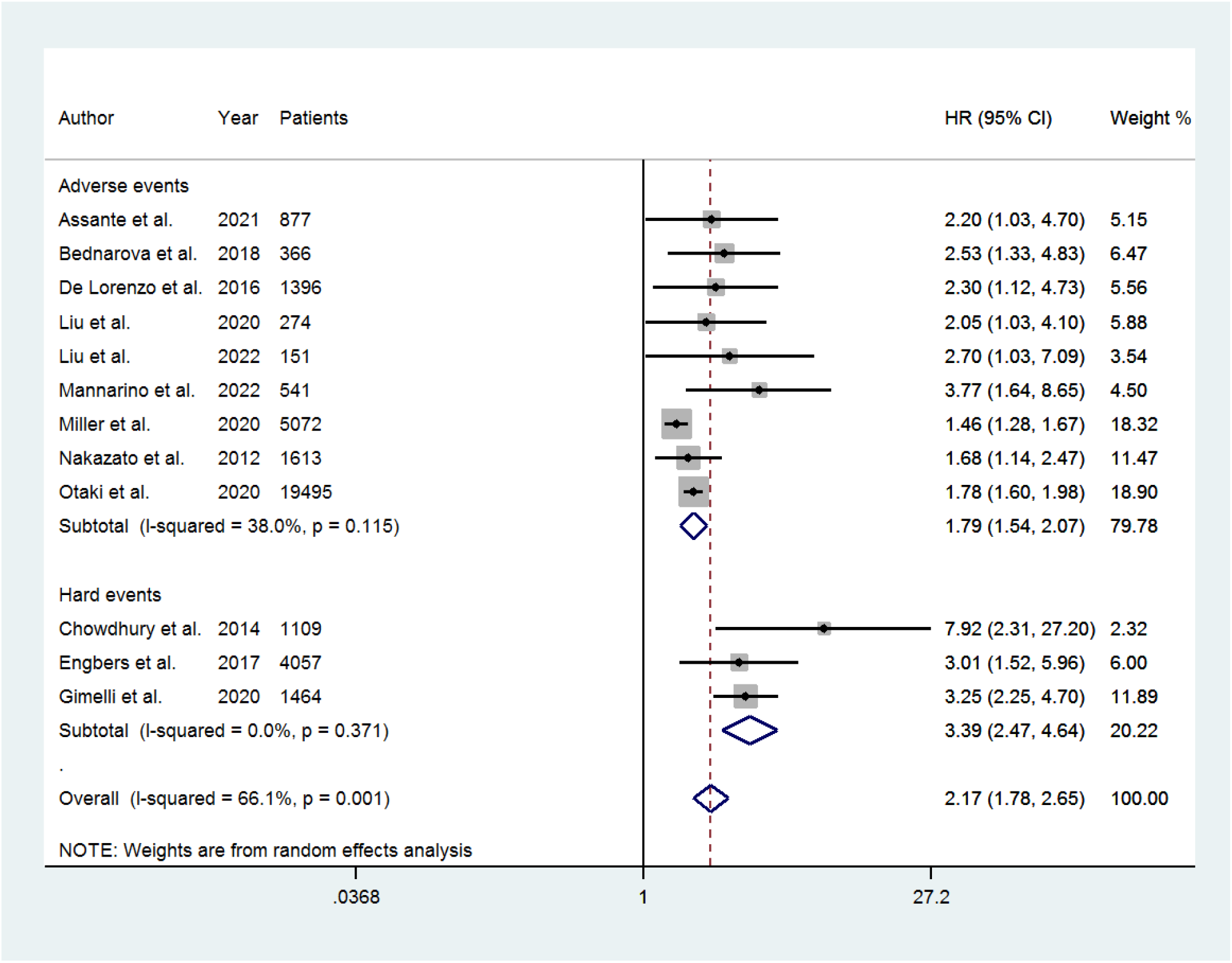
Forest plot of HR for adverse and hard events associated with abnormal myocardial perfusion. Horizontal lines represent 95% CI of the point estimates. The diamond represents the pooled estimate (size of the diamond = 95% CI). The solid vertical line represents the reference of no increased risk, and the dashed vertical line represents the overall point estimate. HR, hazard ratio; CI, confidence intervals.

### Potential Bias and Meta-regression Analysis

The funnel plots showed asymmetry (*P*=0.01) among studies that evaluated adverse events (Figure S2). After applying the trim and fill procedure, the summary random effects HR for adverse events was 1.71 (95% CI, 1.40-2.10) for HR, and the Egger test was no longer statistically significant (*P*=0.81) (Figure S3). At meta-regression analysis we found an association between HR for adverse events and presence of angina symptoms (coefficient 0.009; standard error 0.002; t 3.61; *P*=0.007) and family history of CAD (coefficient 0.019; standard error 0.004; t 4.53; *P*=0.006).

### Subgroups Analysis

A total of five studies reported data on adjusted HR for the occurrence of adverse events. The pooled HR was 1.69 (95% CI, 1.44-1.98) and the heterogeneity was 44.9% (*P*=0.12) (Figure 4). A total of seven studies reported data on unadjusted HR for the occurrence of adverse events. The pooled HR was 2.72 (95% CI, 2.00-3.70) (Figure 4). A total of six studies evaluated patients using D-SPECT camera (Spectrum Dynamics Medical, Haifa, Israel). The pooled HR was 1.82 (95% CI, 1.41-2.34) and the heterogeneity was 37.8% (*P*=0.15) (Figure 5). A total of five studies evaluated patients using Discovery camera (GE Healthcare, Milwaukee, WI, USA). The pooled HR was 3.07 (95% CI, 2.36-3.99) and the heterogeneity was 0.0% (*P*=0.51) (Figure 5).

**Figure 4.**
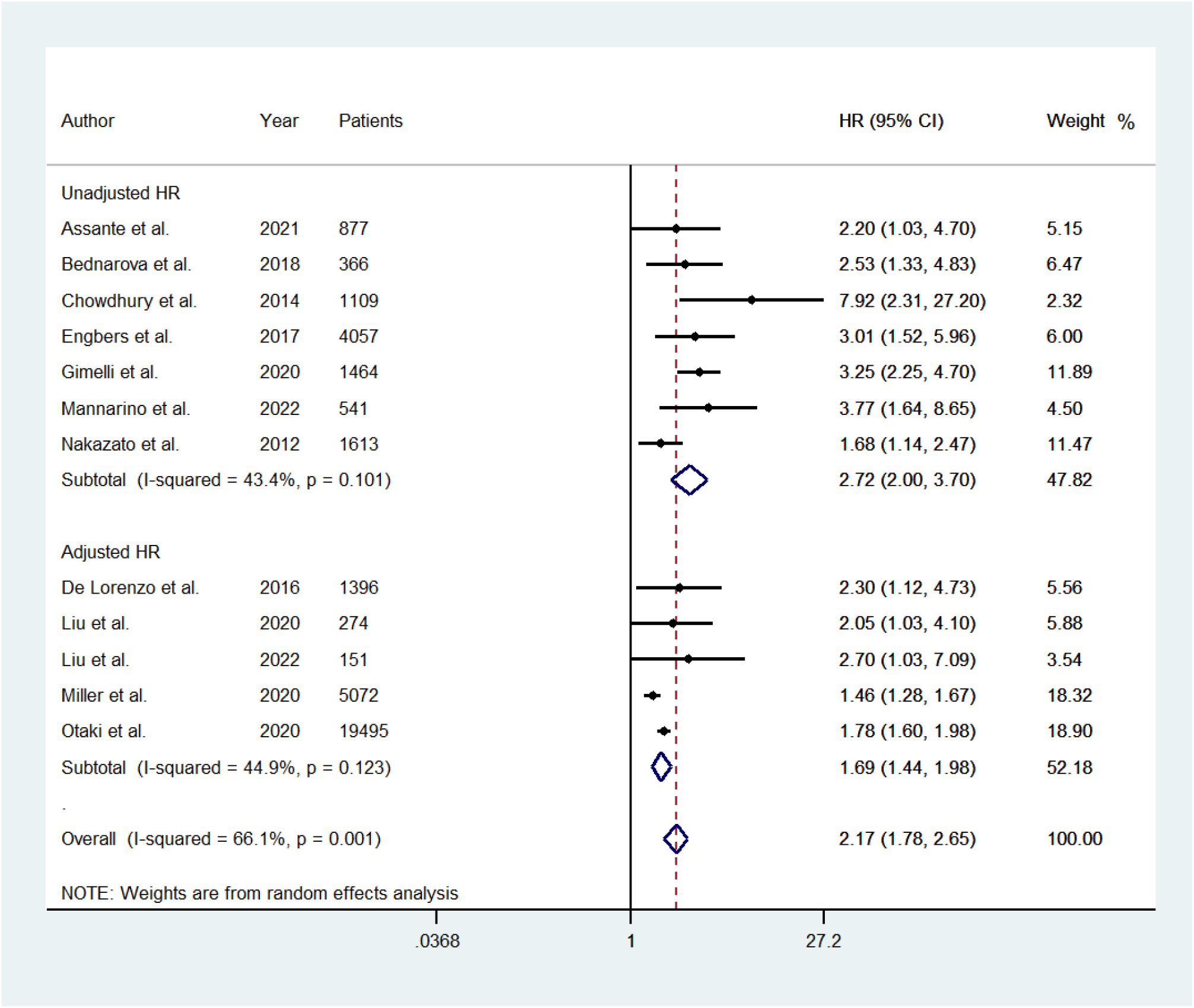
Forest plot for the occurrence of adverse events associated with abnormal myocardial perfusion between studies reporting data on adjusted and unadjusted HR. Horizontal lines represent 95% CI of the point estimates. The diamond represents the pooled estimate (size of the diamond = 95% CI). The solid vertical line represents the reference of no increased risk, and the dashed vertical line represents the overall point estimate. HR, hazard ratio; CI, confidence intervals.

**Figure 5.**
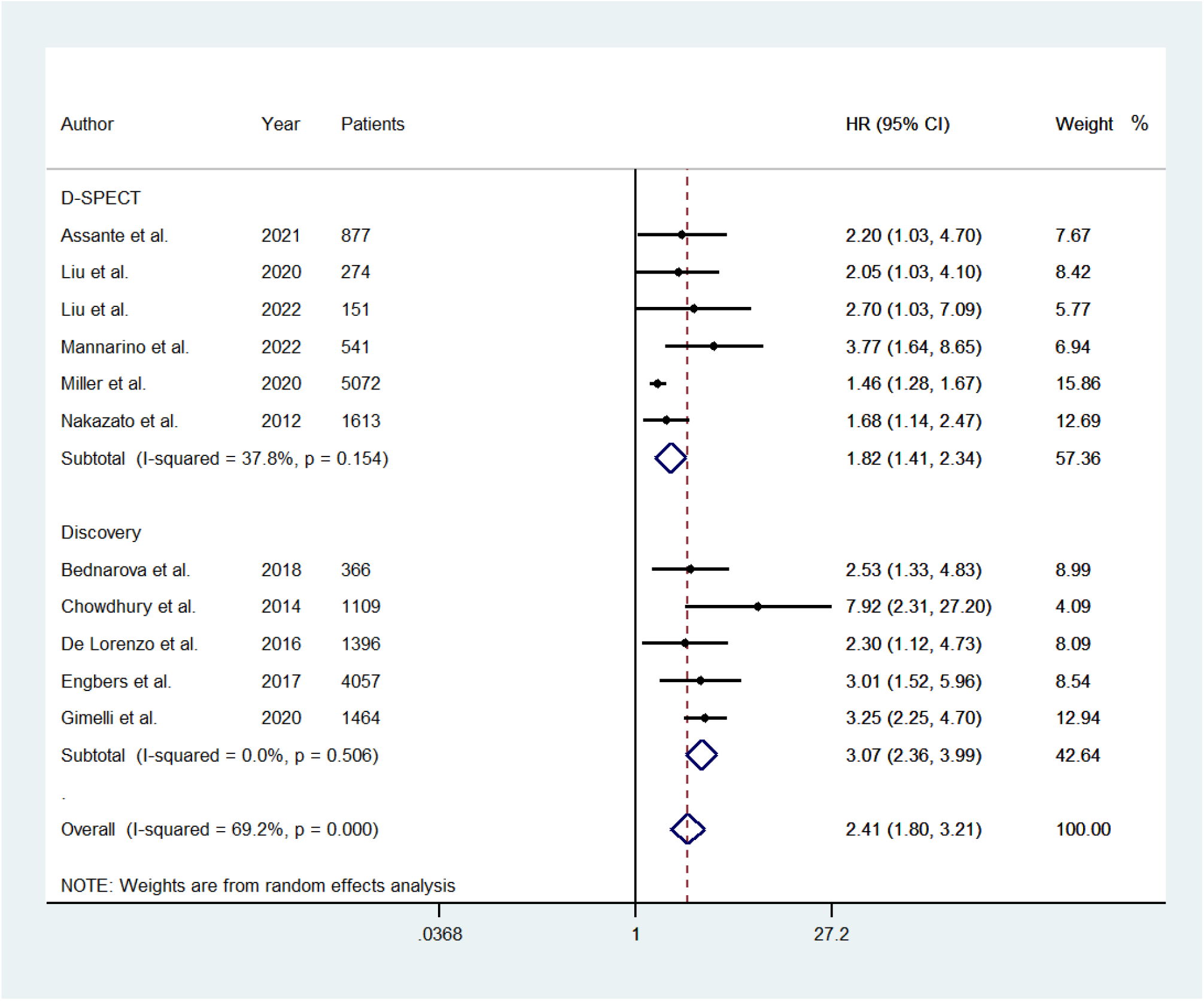
Forest plot of HR of adverse events associated with abnormal myocardial perfusion considering studies using D-SPECT and Discovery camera, respectively. Horizontal lines represent 95% CI of the point estimates. The diamond represents the pooled estimate (size of the diamond = 95% CI). The solid vertical line represents the reference of no increased risk, and the dashed vertical line represents the overall point estimate. HR, hazard ratio; CI, confidence intervals.

### Pooled IRR

A total of nine studies reported data useful to calculate separately the incidence rate of adverse events in patients with abnormal and normal myocardial perfusion imaging. The pooled IRR was 2.38 (95% CI, 1.39-4.06) and the heterogeneity was 84.6% (*P*<0.001) (Figure 6). The funnel plot showed no evidence of asymmetry (*P*=0.52) (Figure S4).

**Figure 6.**
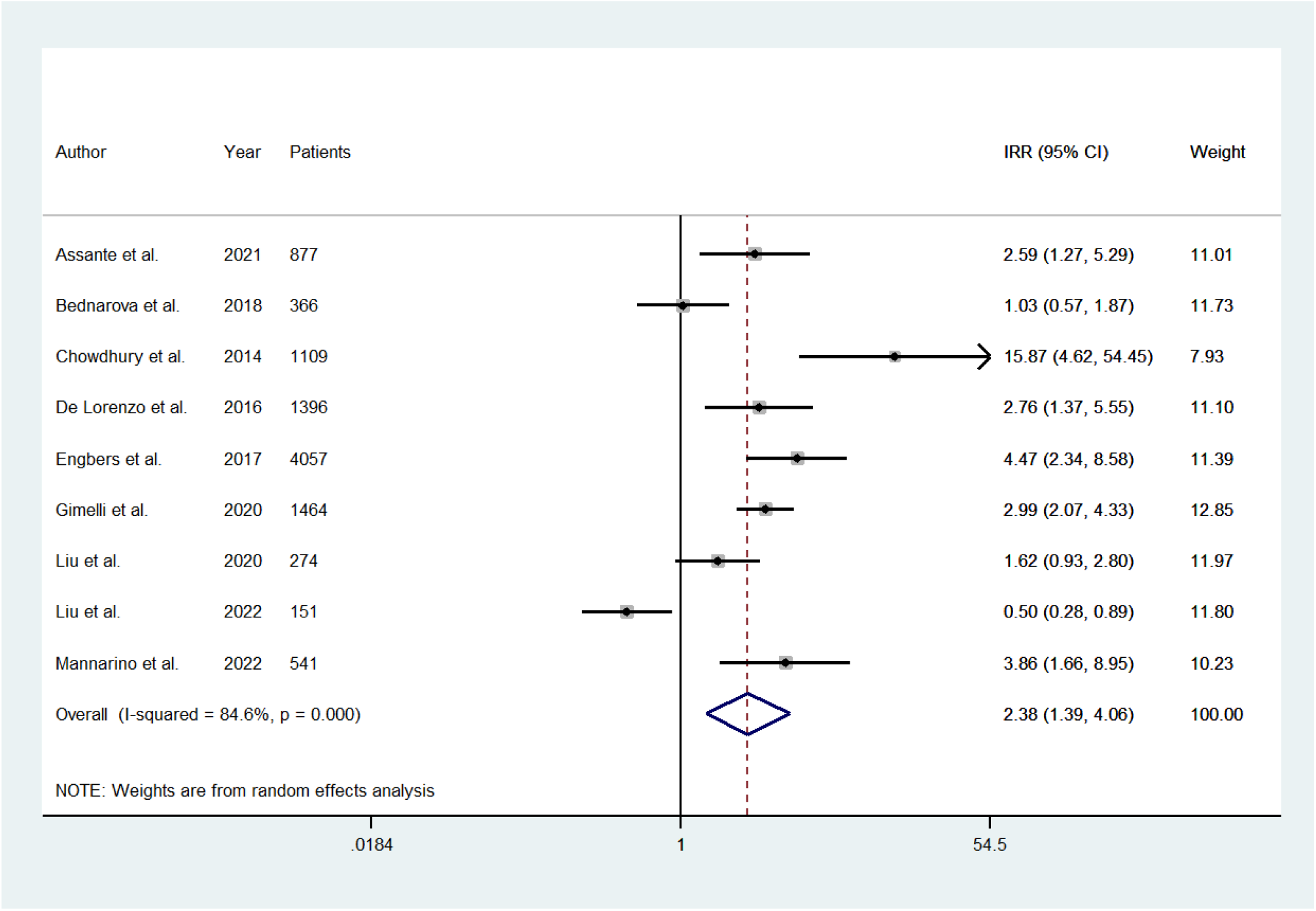
Forest plot for the IRR of adverse events in patients with abnormal and normal myocardial perfusion. lnIRR, natural logarithm of incidence rate ratio; CI, confidence interval.

### Leave-One-Out Sensitivity Analysis

At leave-one-out sensitivity analysis, the summary HR for adverse events (Table S2) remained stable after iteratively removing one study at a time, as well as for the summary IRR of adverse events (Table S3), indicating that our results were not driven by any single study.

## DISCUSSION

To our knowledge, this is the first meta-analysis evaluating the prognostic value of CZT-SPECT in predicting adverse cardiovascular events in patients with suspected or known CAD. By including 12 studies, we found that the pooled estimates of HR for the occurrence of cardiac events is high in the presence of abnormal myocardial perfusion, supporting the evidence on an association between myocardial perfusion abnormalities and adverse outcome. The summary HR of 2.17 for adverse events obtained, indicates that a randomly selected patient with abnormal perfusion has a 68% chance to experience an event before a randomly selected patients with normal perfusion. An abnormal result was associated with a higher probability of events also when the three studies considering only hard events as end point were evaluated.

Stress myocardial perfusion imaging has been widely validated as prognostic predictor of adverse events both in patients with suspected and known CAD during both short-and long-term follow-up,^37–39^ showing to be cost-effectiveness also in comparison with other imaging modalities.^40^ As reported different years ago in different peer-reviewed articles in overall 69,655 patients, there is a clear separation in risk of major adverse cardiac events between patients with low- and high-risk perfusion imaging results over the ensuing 2 to 4 years of follow-up.^39^ In particular, by using traditional Anger camera, prior published data showed the clear association between the extent and severity of myocardial perfusion variables and the risk of cardiac events.^39^

The use of CZT camera has increased in the last decade, thanks to some advantages compared to standard Anger camera. In particular, the higher sensitivity and related reduced acquisition time have led to an optimal diagnostic accuracy, with better patient compliance as compared to traditional camera.^41^ A previous meta-analysis demonstrated that despite conventional Anger camera and CZT-SPECT have both good diagnostic performance in detecting CAD, CZT-SPECT showed a slightly higher accuracy, supporting the use of this system in clinical practices.^41^

Since its introduction, several studies have pointed up on diagnostic and prognostic value of CZT-SPECT in the evaluation of patients with suspected or known CAD.^9–11, 42^ Published data found that the prognostic value of CZT-SPECT was similar or slightly higher compared to conventional Anger camera in predicting adverse outcome.^9–11^ In particular, using lower radiation dose and shorter acquisition time by CZT camera, an optimal risk stratification has been obtained, with a lower prevalence of adverse events in patients with normal scan.^9^ It has been pointed out that normal perfusion by CZT is associated with a lower rate of adverse cardiac events.^34^ Moreover, similarly to results obtained by using conventional camera, all the studies using CZT confirmed that the extent of abnormality was associated with higher risk of events, and patients with abnormal findings had a significantly higher risk during follow-up.

The recent multicenter REgistry of Fast Myocardial Perfusion Imaging with NExt generation SPECT (REFINE)^43^ an observational study collecting clinical, imaging and prognostic data from 20,418 patients from 5 institutions, using different CZT models and protocols, confirmed the relationship between perfusion abnormalities and prognosis. In particular, total perfusion defect (TPD), provided an excellent power in risk stratification.^44^ Interestingly, also in patients with more than 0 but less than 1% TPD had higher risk of events than patients with TPD 0. This aspect has been correlated with the higher spatial resolution of CZT camera.

Based on data from 36,415 patients derived from 12 independent cohorts, we showed that the presence of an abnormal finding conferred a 2-fold increased risk of cardiovascular events. Our meta-analysis demonstrates consistently that the presence of abnormal findings may identify patients who warrant more intensive clinical surveillance for adverse cardiac events.

In the present meta-analysis, most of the selected studies used the summed stress score to assess the presence of abnormal perfusion, while only two studies evaluated the presence of ischemia as predictor of events. It should be considered that although the summed stress score is the perfusion imaging variable most extensively validated for cardiac risk stratification, the summed difference score represents the variable most predictive of subsequent coronary angiography and early revascularization. Thus, further studies are needed to better define the role of the extent and severity of myocardial ischemia by CZT in clinical decision making and prognostic evaluation. The estimated IRR of adverse events in patients with abnormal and normal myocardial perfusion by CZT-SPECT confirmed that those with abnormal perfusion had a worse outcome. In a subgroup analysis, evaluating only data on adjusted or unadjusted HR and considering studies by two different CZT cameras, D-SPECT or Discovery, the results on prognostic value were confirmed, without heterogeneity.

### Strengths and Limitations

It should be underlined that the excellent prognostic value of CZT camera can be reached by using lower radiation dose and shorter acquisition imaging time compared to standard camera. These advantages should also be implemented by stress-first imaging protocol, considering that CZT camera imaging findings are more often interpreted as normal. This aspect is relevant as the problem of radiation exposure and related hazard, and the awareness that imaging modality needs to improve clinical outcome, could led to choose an imaging modality with lower radiation dose associated to a validated clinical benefit for patients.^45^

A limitation of our meta-analysis is that we analyzed a dichotomous criterion for imaging abnormality (abnormal vs. normal findings) for CZT-SPECT studies. Available literature supports the concept that semi-quantitative SPECT scoring, including the extent and severity of myocardial perfusion defects, are highly predictive of patient outcome. However, our chose was made to limit the heterogeneity of available studies, due to the differences in abnormal SPECT finding classification. It is possible that a more quantitative index of extent and severity would have increased the predictive power of CZT-SPECT. Moreover, differences in distribution of patients with known or suspected CAD among included studies and differences in stress test, acquisition protocol and the variety of endpoint considered might impact the predictive value of CZT-SPECT. Hence, potential bias should be adequately considered in interpreting the observed results. Finally, individual outcome data were not available in the studies included in the meta-analysis.

### Conclusions

The results of this meta-analysis demonstrate that an abnormal myocardial perfusion at stress CZT-SPECT is associated with adverse cardiovascular events. Our study underlines that stress myocardial perfusion imaging by CZT-SPECT represents a valuable noninvasive prognostic indicator for cardiac events in patients with known or suspected CAD.

## Data Availability

All the data in the manuscript are available

## Sources of Funding

None.

## Disclosures

None.

## Supplemental Material

Detailed PubMed search strategy

PRISMA checklist

Figures S1-S4 Table S1-S3

Supplementary References

## Nonstandard Abbreviations and Acronyms

CAD: Coronary artery disease
CZT: Cadmium-zinc-telluride
HR: Hazard ratio
IRR: Incidence rate ratio
SPECT: Single photon emission computed tomography
TPD: Total perfusion defect

## Notes

### Competing Interest Statement

The authors have declared no competing interest.

### Funding Statement

No funding

### Author Declarations

CE Protocol number 110/17

